# Usefulness of real-time RT-PCR to understand the kinetics of SARS-CoV-2 in blood: a prospective study

**DOI:** 10.1101/2022.03.07.22271764

**Authors:** Nelly Daniela Zurita-Cruz, Alexandra Martín-Ramírez, Diego Aníbal Rodríguez-Serrano, Isidoro González-Álvaro, Emilia Roy-Vallejo, Rafael De la Cámara, Leticia Fontán García-Rodrigo, Laura Cardeñoso Domingo

## Abstract

**Background:** SARS-CoV-2 viral load and kinetics assessed in serial blood samples from hospitalised COVID-19 patients by RT-PCR are poorly understood.

**Methods:** We conducted an observational, prospective case series study in hospitalised COVID-19 patients. Clinical outcome data (Intensive Care Unit admission and mortality) were collected from all patients until discharge. Viremia was determined longitudinally during hospitalisation, in plasma and serum samples using two commercial and standardised RT-PCR techniques approved for use in diagnosis of SARS-CoV-2. Viral load (copies/mL and log10) was determined with quantitative TaqPath™COVID-19 test.

**Results:** SARS-CoV-2 viremia was studied in 57 hospitalised COVID-19 patients. Persistent viremia (PV) was defined as two or more quantifiable viral loads detected in blood samples (plasma/serum) during hospitalisation. PV was detected in 16 (28%) patients. All of them, except for one who rapidly progressed to death, cleared viremia during hospitalisation. Poor clinical outcome occurred in 62.5% of patients with PV, while none of the negative patients or those with sporadic viremia presented this outcome (p<0.0001). Viral load was significantly higher in patients with PV than in those with Sporadic Viremia (p<0.05). Patients presented PV for a short period of time: median time from admission was 5 days (Range=2-12) and 4.5 days (Range=2-8) for plasma and serum samples, respectively. Similar results were obtained with all RT-PCR assays for both types of samples.

**Conclusions:** Detection of persistent SARS-CoV-2 viremia, by real time RT-PCR, expressed as viral load over time, could allow identifying hospitalised COVID-19 patients at risk of poor clinical outcome.

**Highlights:**

- Commercial RT-PCR techniques could be used to monitor SARS-CoV-2 viremia kinetics.
- SARS-CoV-2 persistent viremia is related with poor outcome in COVID-19 patient.
- SARS-Cov-2 viremia kinetics could be used as a biomarker of poor prognosis.
- Plasma samples are the best choice for analysis of SARS-CoV-2 viremia kinetics.

## INTRODUCTION

Severe Acute Respiratory Syndrome Coronavirus-2 (SARS-CoV-2) was first described in December 2019 in Wuhan, China (1). As of 19th of January 2022, 5,542,359 deaths were reported to WHO(2).

While most COVID-19 patients present mild disease, others develop a severe disease (3). Identifying patients at risk of developing severe COVID-19 is an unmet need to improve the management of these patients and prevent morbidity and mortality. Some risk factors of poor prognosis, such as elevated concentrations of interleukin (IL)-6, contribute to risk stratification of COVID-19 patients (4–6).

Detection of SARS-CoV-2 RNA by real-time reverse transcription polymerase chain reaction (RT-PCR) in nasopharyngeal swabs is the gold standard for COVID-19 diagnosis (7). However, no consistent association has been reported between the viral load in these samples and disease severity (8).

Recently, detection of SARS-CoV-2 RNA in blood samples (viremia) has been considered as a potential predictor of poor prognosis, due to its association with rapid deterioration and death (9–13). Nevertheless, there is a lack of reports analysing viral load quantification over time in sequential samples, which could allow understanding viremia kinetics and its relationship with the patient clinical outcome.

The objectives of this prospective study were to analyse the kinetics of SARS-CoV-2 viremia, by qualitative and quantitative RT-PCR methods using standardised commercial reagents in sequential samples collected longitudinally from COVID-19 patients during hospital admission, and to determine its relationship with disease clinical course.

## MATERIAL AND METHODS

### Study design and patients

This is a prospective longitudinal case series study, conducted at Hospital Universitario de La Princesa, Madrid (Spain), from November 2020 to January 2021. Fifty-seven consecutively admitted patients with COVID-19 diagnosed by RT-PCR from nasopharyngeal swabs were included. All patients gave their informed consent, and the research protocol was approved by the Institutional Ethics Review Board (register number 4267). We followed guidance from The Strengthening the Reporting of Observational Studies in Epidemiology (STROBE) standards for observational research, and those of the Updated List of Essential Items for Reporting Diagnostic Accuracy Studies (STARD) (14).

### Data collection and definitions

In addition to sociodemographic variables, clinical characteristics and outcome variables (Intensive Care Unit [ICU] admission and mortality during hospitalisation) were collected. “Poor Outcome” was considered when the patient had at least one of the two clinical variables described above. Other relevant variables collected were symptom onset, hospital admission, hospital discharge, death and sample collection dates. All data collected was entered in an anonymized database with access only to the research team.

### Sample collection

First, a blood sample (serum and plasma) was collected within the first 24-36 hours upon admission; subsequently, additional blood samples were collected every 48-72 hours during the first week. Later, samples were collected twice a week until discharge. Samples were frozen at - 80ºC until RT-PCR performance.

### Qualitative RT-PCR Methods

All blood samples were assessed by two qualitative RT-PCR methods: Cobas®SARS-CoV-2 Test, Roche Diagnostics (cobas®-test), an automated method for SARS-CoV-2 detection; and TaqPath™COVID-19 CE IVD RT-PCR Kit, Thermo Fisher Scientific (TaqPath™-test), according to manufacturers’ indications (details are shown in supplementary material). For qualitative detection, samples were assessed in duplicate by TaqPath™-test and only once by cobas®-test due to scarce volume.

### Quantification of SARS-CoV-2 viral load

Quantitative RT-PCR to determine viral load (qTaqPath-test) was performed using TaqPath™-test reagents in a QuantStudio™ 5 Real Time PCR System (Applied Biosystems). Plasma and serum samples were analysed in duplicate. Viral load quantification of serum and plasma samples was obtained by plotting Ct values through the standard curve obtained using the QuantStudio™ Design and Analysis software version 2.4.3 (Applied Biosystems, USA) and according to Thermocycler manufacturer specifications (15). Detailed description of standard curve determination is shown in supplementary material. Samples were considered quantifiable when mean Ct in the duplicate test for each gene was ≤37 and standard deviation (SD) was <0.5. All results not fulfilling these criteria and/or those with detection in only one duplicate, were considered positive, but not quantifiable. To estimate a value for quantification limit, median of all non-quantifiable values was calculated, obtaining 17.75 copies/mL for serum samples and 14.89 copies/mL for plasma samples. For the genes that met quantification criteria, mean quantity was calculated from the duplicates, expressed as copies/mL and logarithm with base 10(log10). Two positive controls (corresponding to 20,000 and 200 copies) and two negative controls were added in each run in duplicate.

Moreover, we studied the number of genes (N, S or ORF) that were amplified in each plasma sample tested by q-RT-PCR, as an indication of the association of viremia with the presence of virions.

### Kinetics of SARS-CoV-2 in blood

The presence of SARS-CoV-2 RNA was detected by qualitative RT-PCR methods and viral load was determined by qTaqPath-test in plasma and serum of each patient collected throughout the follow-up. To analyse viremia kinetics, time course curves were obtained plotting viral load change over time. Specific patterns of viral load change were identified and their relationship with clinical evolution was analysed.

Statistical analysis is detailed in supplementary methods.

## RESULTS

### Characteristics of the study population

Fifty-seven patients consecutively admitted to our hospital were included in this study (Figure 1). A median of 4 samples per patient (R=2-18) was collected. Median age was 64 years (R=31-94 years) and 61.4% were male. During hospital admission, 8 (14.0%) patients required ICU admission (median age 59.5 years; R=52-76 years; 87.5% male); and 5 (9%) died during hospitalisation (median age 76; R=59-86 years; 80% male). Some patients presented more than one clinical condition. Patients included in the Poor Outcome group (n=10; 17.5%) had a median age of 60.5 years; R=52-86 years, 85.7% were male.

**Figure 1:**
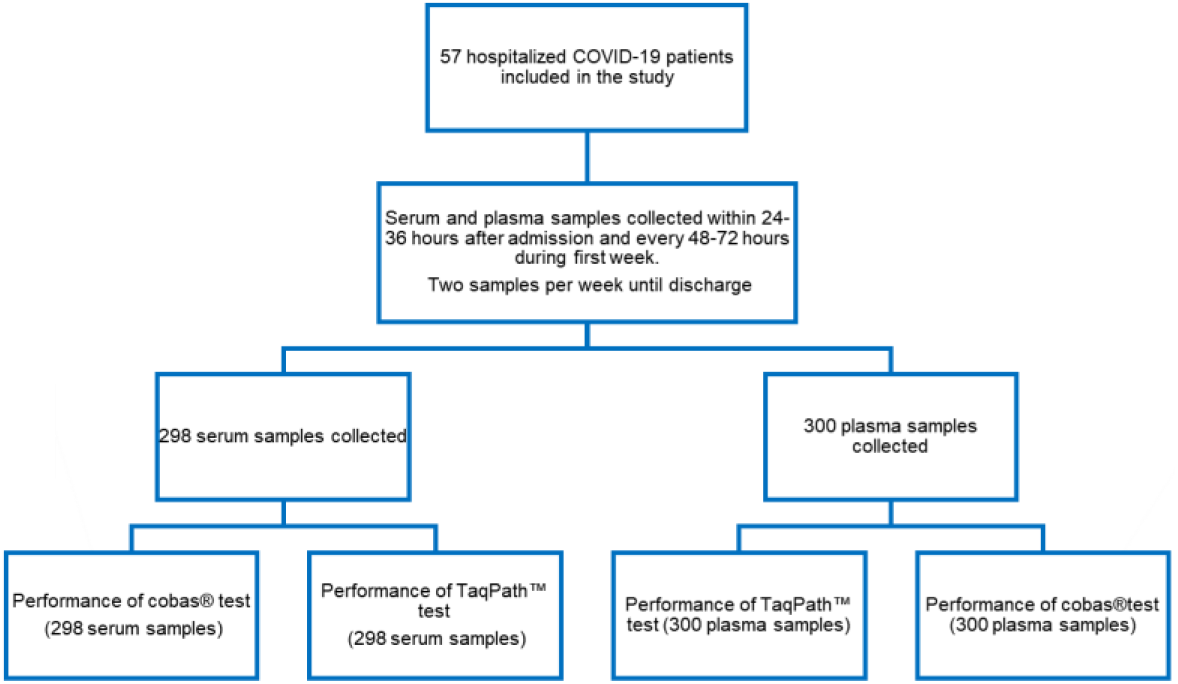
STROBE flow chart of the study including participation, sample collection and tests.

### Analysis of samples and RT-PCR techniques

A total of 598 samples (298 serum and 300 plasma samples) were assessed (Figure 1).

Qualitative detection obtained by cobas® and TaqPath™ tests showed a similar concordance (κ=0.613 and κ=0.604, respectively) for both types of samples. However, both tests showed statistically significant higher positive detection in plasma samples (Table 1).

**Table 1.** Agreement of viremia qualitative detection in serum and plasma samples according to RT-PCR technique.

|  | cobas®-test (n=298) |  | TaqPath™-test (n=300) |  |
| --- | --- | --- | --- | --- |
|  | Plasma Positive n (%) | Plasma Negative n (%) | Plasma Positive n (%) | Plasma Negative n (%) |
| **Serum Positive n (%)** | 75 (25.2) | 17 (6) | 76 (25.3) | 13 (4.6) |
| **Serum Negative n (%)** | 35 (11.7) | 171 (57.8) | 41 (13.3) | 170 (56.7) |
| **Agreement** | 82.9% |  | 82% |  |
| **Kappa coefficient** | 0.613 |  | 0.604 |  |
| **p value** | <0.0001 |  | <0.0001 |  |

Mean Ct values obtained from plasma and serum by TaqPath™-test (33.99±2.05 and 34.21±1.94, respectively) and those obtained by cobas®-test (35.34±2.12 and 35.16±2.33, respectively) did not show significant differences (p>0.5). Pearson correlation analysis of mean Ct showed correlation coefficients (r) of 0.86 and 0.76 for plasma and serum samples, respectively (Figure 2). Results of the correlation of Ct obtained by both techniques in serum samples are consistent with findings described in our previous series(16).

**Figure 2:**
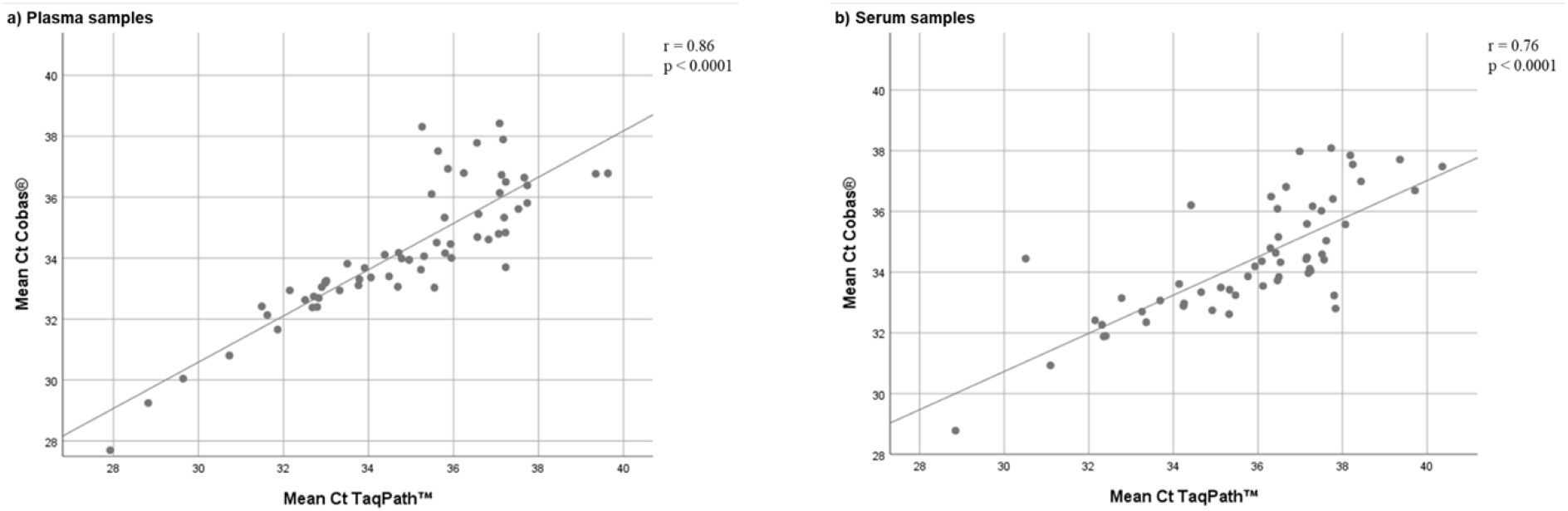
Scatter plot of mean Ct obtained by Taqpath™ and cobas® RT-PCR tests in plasma (a) and serum (b). Correlation between results was analysed by Pearson correlation (r=0.86 and 0.76, respectively. P<0.0001 for both techniques).

### Analysis of SARS-CoV-2 viral loads

Viral load could be quantified with qTaqPath-test in 50 plasma and 41 serum samples. Median viral load was 462.88 copies/mL (R=37.26-21,277) for plasma, and 370 copies/mL (R=29.42-15,257) for serum samples. Viral load expressed in log10 showed a normal distribution. Mean value of viral load in plasma was 2.65±0.63 log10 and 2.6±0.58 log10 for serum samples. No statistically significant differences were found between both types of samples (p=0.65). Furthermore, correlation analysis of viral load obtained from plasma and serum showed a Pearson correlation coefficient r=0.89 (Figure 3).

**Figure 3:**
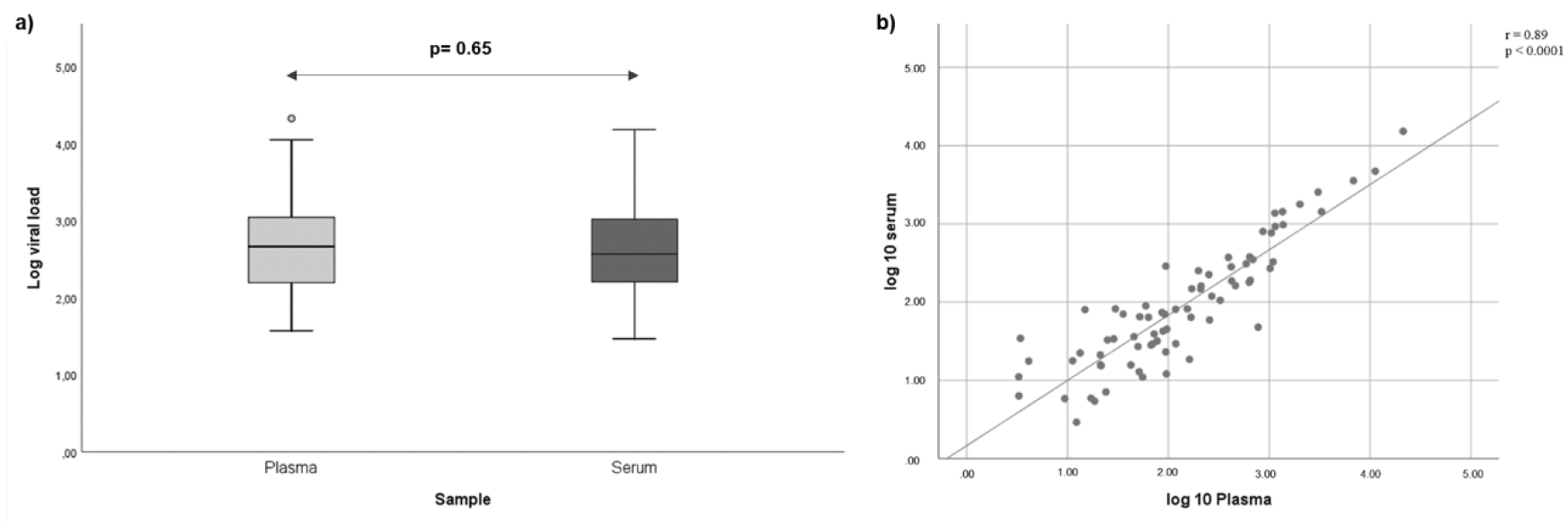
Comparison of viral load in plasma and serum samples. Viral loads were determined by qTaqPath-test. Differences between plasma and serum were analysed by Student’s t test (p=0.65) (a). Correlation between data was evaluated by Pearson correlation coefficient (r=0.89; p<0.0001) (b).

### Detection of SARS-CoV-2 genes in plasma samples

Detection of SARS-CoV-2 genes in plasma by TaqPath™ Kit showed that at least two assay targets could be detected in 79% of the patients: all 3 targets were detected in 64.3%, 2 targets in 14,3% and only the N gene in 21.4% of the cases. (Figure 4).

**Figure 4.**
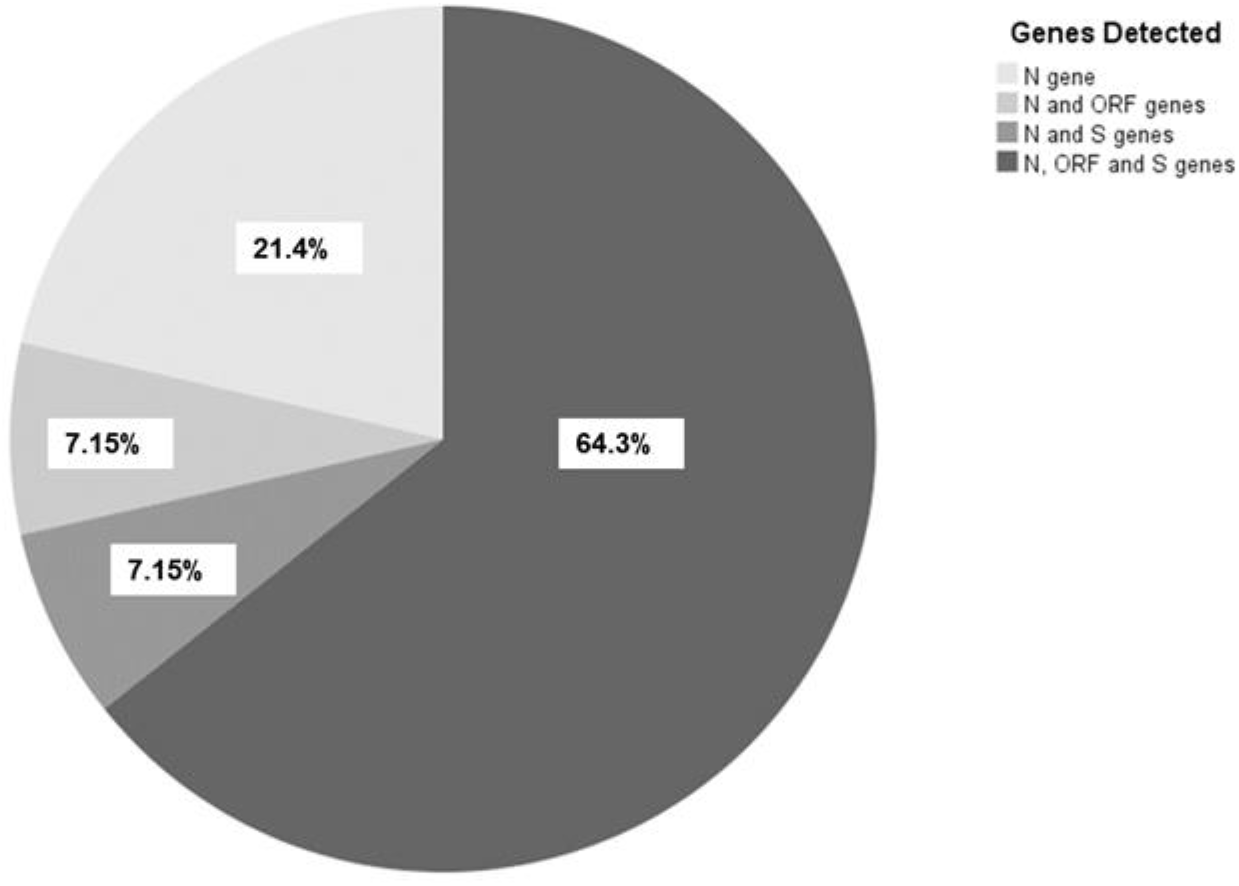
Distribution of genes detection by TaqPath™ kit in patients with positive viremia.

### Kinetics of SARS-CoV-2 viremia in hospitalised patients

Viremia analysis over time was performed in consecutive plasma and serum samples of individual patients (n=57) through the qualitative methods, cobas® and TaqPath™ tests, and viral load was assessed using qTaqPath-test. Three different viremia patterns were identified: i) Persistent Viremia (PV), viremia was detected in two or more consecutive determinations, n=16 (28%); ii) Sporadic Viremia (SV), viremia was only detected in isolated samples, n=34 (60%); and, iii) Negative Viremia (NV), viremia was not detected in any of the samples during hospitalisation, n=7 (12%).

Patients presented PV for a short period of time: median time was 5 days (Range=2-12) for plasma samples and 4.5 days (Range=2-8) for serum. Viremia clearance occurred spontaneously in PV patients during hospitalisation, except for one patient who progressed rapidly to death.

Analysis of viral load in consecutive samples of all patients with PV (n=16) showed different trends. Nine (56.2%) patients showed a parabolic curve, with an initial increase of viral load followed by a decrease. Six (37.5%) patients, 5 of them admitted to ICU, showed a decreasing viral load curve. Conversely, one patient (6.3%) showed increasing viral load until decease. (Figure 5)

**Figure 5.**
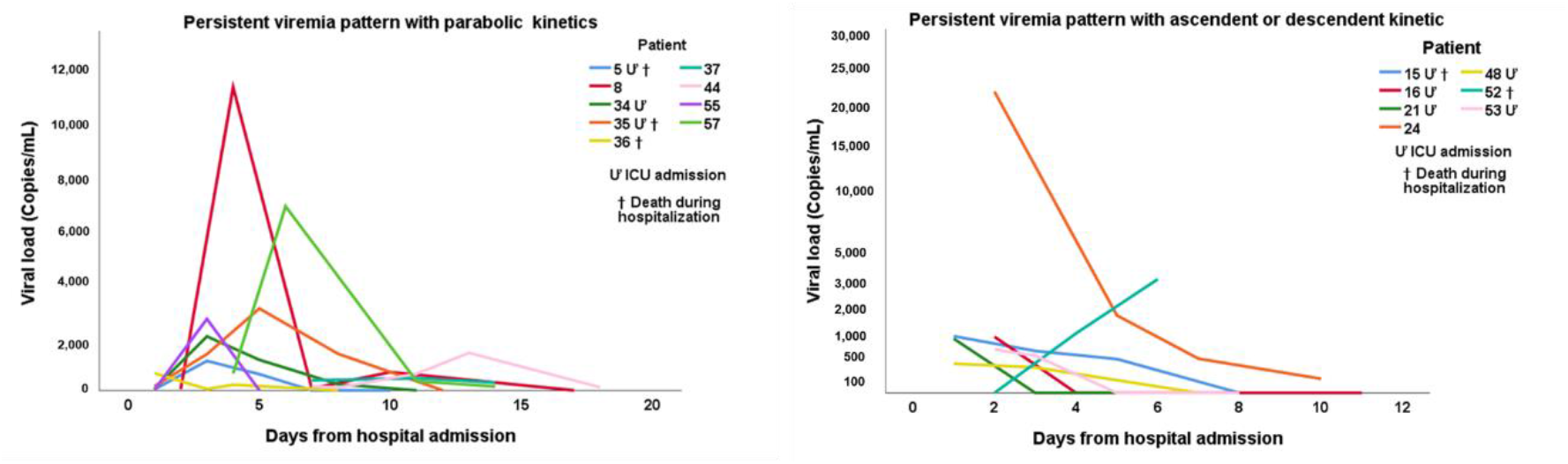
Viral load kinetics curves throughout hospitalisation in plasma from patients with Persistent Viremia. Clinical characteristics defining poor outcome are included within the graphics for each patient. ICU: intensive care unit.

To further characterise the kinetics of SARS-CoV-2 in the blood of patients with PV, both viremias assessed by the 2 qualitative techniques (expressed a Ct) and viral load assessed by the q-TaqPath test (expressed as copies/mL) were determined in plasma and serum samples. Kinetics curves represent the change of these parameters in individual PV patients during hospitalisation for the qualitative (Figure 6A) and quantitative (Figure 6B) determinations. Curves obtained for each individual patient showed similar kinetic behaviour regardless of the technique or the type of sample used for the analysis. These results suggest that PV could be monitored using plasma or serum samples and both, qualitative and quantitative RT-PCR methods.

**Figure 6.**
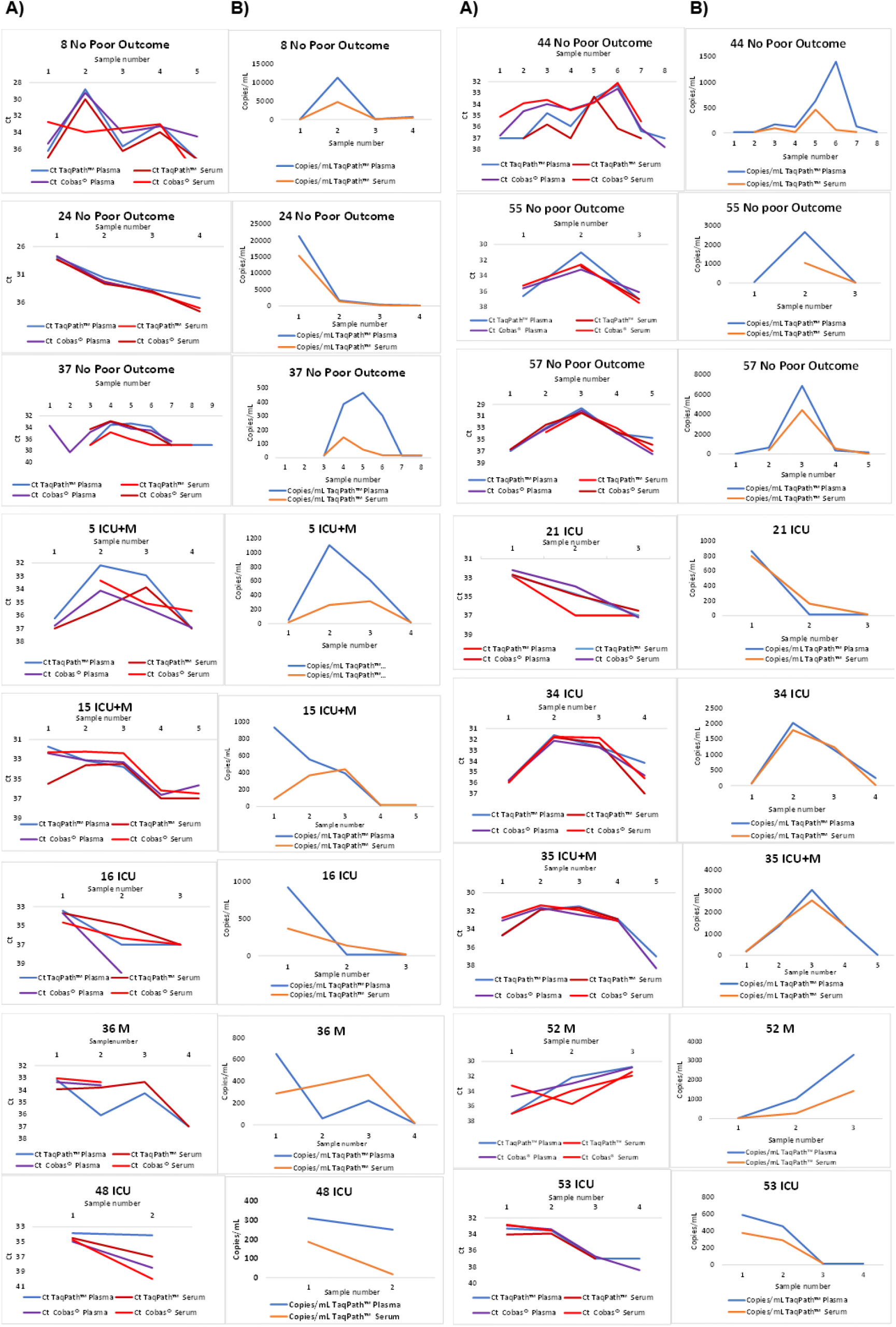
Analysis of SARS-CoV-2 viremia kinetics assessed by different RT-PCR methods in plasma and serum samples. The headings of the graphs represent the code of the anonymized patients, followed by their clinical outcome (ICU: Intensive Care Unit admission; M: Death during hospital admission). The curves show the kinetics of viremia determined by the TaqPath™-test and cobas®-test, expressed as Ct values (A), and the q-TaqPath test, expressed in copies/mL (B).

### Relationship of Persistent Viremia with clinical outcome and viral load in blood

Analysis of viral load in consecutive samples over time and its relationship with clinical outcome showed that 100% of those patients with Poor Outcome (ICU admission and/or death) during hospitalisation (N=10) presented PV, whereas none of those patients with SV and NV presented a Poor Outcome and all of them were discharged. Although 6 PV patients did not present any characteristic of poor clinical outcome, those patients represented only the 13% of all patients without Poor Outcome (N=47), that is, poor clinical outcome was significantly associated with PV compared to no PV (p <0.0001), as shown in Figure 7.

**Figure 7.**
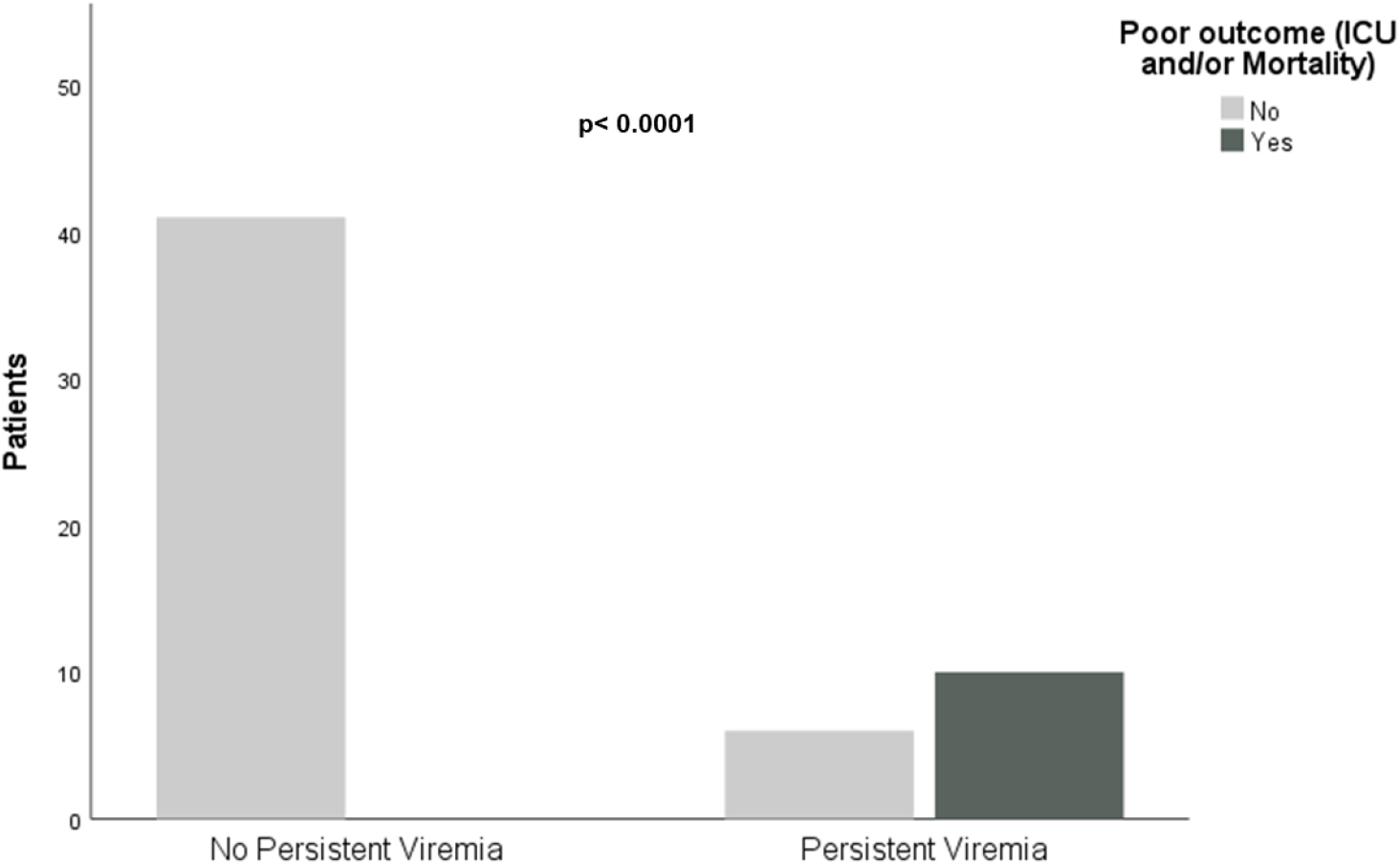
Relationship of Persistent Viremia with clinical outcome. Distribution of patients according to the presence or absence of Persistent Viremia is shown. The comparison between both groups was analysed with the χ2 test.

No difference was detected in age (p=0.2) or sex (p=0.4) between patients with different viremia patterns.

The viral load had a normal distribution when it was expressed as log10. Regarding viral load in patients with different viremia patterns, SV patients showed lower viral loads than PV. Median viral loads for SV were 77.3 copies/mL (IQR= 37.3-77.3; mean= 1.82±0.22 log10) and 59.4 copies/mL (IQR=29.4-59.4; mean=1.71±0.34 log10) for plasma and serum, respectively, whereas for PV median viral load was 558 copies/mL (IQR= 170.3-1145.4; mean= 2.71±0.61 log10) for plasma and 370.4 copies/mL (IQR= 180.92-1233.6; mean=2.57±0.55 log10) for serum. Viral load was significantly higher for PV compared to SV for both types of samples (Figure 8).

**Figure 8.**
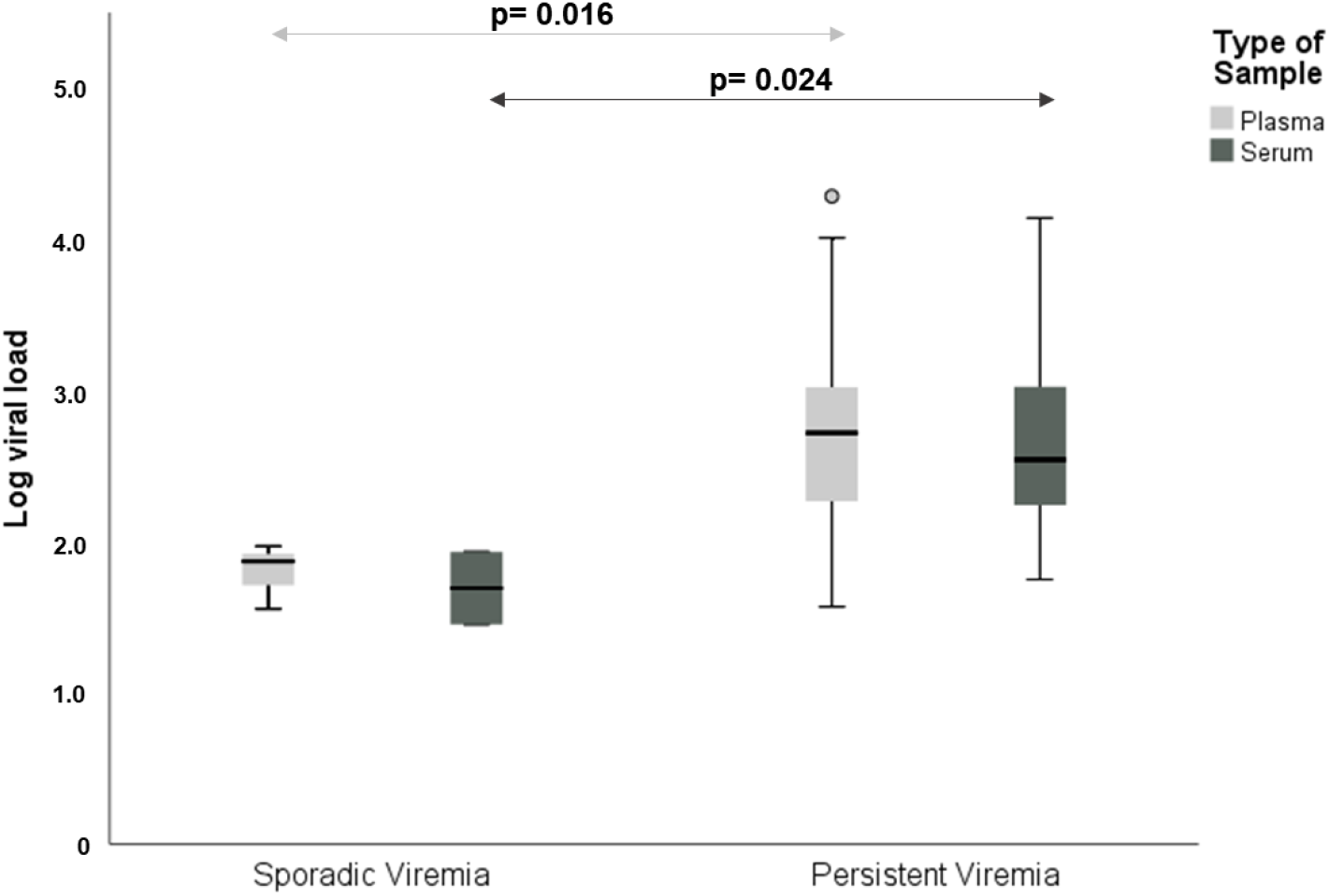
Viral load is significantly higher in patients with persistent viremia than those with sporadic viremia in serum and plasma samples. The viral load expressed as log10 showed a normal distribution, so the difference of viral loads between both groups was analysed by Student’s t test.

## DISCUSSION

The hypothesis of this study was that detectable SARS-CoV-2 viremia in successive samples over time (persistent viremia) could identify hospitalised COVID-19 patients with high risk of poor clinical outcome. To verify this, we studied methods for qualitative detection of SARS-CoV-2 RNA and for quantification of viral load in plasma and serum, using commercially available RT-PCR kits, marked with CE and FDA authorisation. In addition, the association of viremia evolution with patient outcome was also evaluated.

Regarding the relationship between disease severity and the pattern of viremia kinetics, our results showed that all patients with ICU admission and/or death during hospitalisation had PV, while none of the patients with SV or NV had poor clinical outcome in this cohort. This supports our hypothesis that patients with PV are more likely to have a poor clinical outcome, in agreement with other authors. (17–23)

The graphical representation of viremia kinetics patterns in patients with PV, assessed by two qualitative and one quantitative RT-PCR methods showed similar kinetic patterns regardless of the use of different reagents and RT-PCR platforms. This analysis suggests that the dynamics of viremia in longitudinal samples correlates with the presence of SARS-CoV-2 in blood and is reproducible. However, some discrepancy values were detected in isolated cases where the Ct value or viral load was lower than those in the paired sample from the same blood extraction (plasma vs. serum). We hypothesised that those discrepancies could be due to a loss of sample integrity.(24) However, the fact that different techniques can be used to analyse the kinetics of the SARS-CoV-2 viremia increases the versatility of this determination when it comes to its implementation in clinical microbiology laboratories.

Our previous results suggested that the Ct value is a good approximation for the stratification of severely ill patients(13). However, the variation due to the sample type and handling, the technique used, and the intrinsic variability of RT-PCR makes it necessary to develop a standardised method to obtain more reproducible results(25). Even in this series, both assays detected a higher number of positives in plasma than in serum. In this sense, it is well known that the quantification of viral load in plasma has been standardised in other pathologies associated to viral infections such as HIV, HCV, HBV, CMV(26–29), and provides a more precise and reliable monitoring of viremia than Ct values. For this reason, we propose the use of SARS-CoV-2 viral load from plasma samples for the analysis of viremia kinetics.

Different authors have quantified viremia using new technologies such as Droplet Digital PCR (18, 31) using internally developed methods with primer design (17), or ultrasensitive quantitative RT-PCR(32). In this context, the objective of this work was to assess the usefulness of a quantification method using RT-PCR to monitor SARS-CoV-2 viral load in hospitalised COVID-19 patients. We used commercial and standardised reagents with CE and FDA marking for authorisation, making a standard curve and taking advantage of the options offered by the QuantStudio 5 thermal cycler interpretation software. Taken all together, these characteristics provide a robust and reproducible quantification method that allows monitoring viremia in hospitalised COVID-19 patients. Also, we agree with other authors on the need to develop standardised techniques for quantifying viral load in blood, with FDA or CE approval, that could contribute to improve the management of patients with COVID-19(17, 21).

The detection of SARS-CoV-2 RNA in blood has been called RNAemia by some authors (24, 30, 31). However, we refer to this determination as viremia since, in almost 80% of the samples studied in this study, 2 or 3 SARS-CoV-2 genes were detected; the presence of these genes could be correlated with the presence of viral particles (32, 33). In addition, detection of RNA in successive blood samples would indicate the presence of the virus.

The present study has certain limitations. First, the number of patients included in the study was moderate (57 patients). Second, although two RT-PCR techniques were performed, only the TapPath™ test allowed us to quantify the viral load (in copies/ml and log10 viral load).

In summary, we conclude that persistent SARS-CoV-2 viremia in blood samples may be potentially used as indicator of poor prognosis in hospitalised COVID-19 patients. Both qualitative and quantitative RT-PCR techniques are suitable for the characterisation of viremia kinetics in plasma and serum samples. Knowledge of SARS-CoV-2 kinetics in blood allows stratification of hospitalised COVID-19 patients.

## Data Availability

All data produced in the present study are available upon reasonable request to the authors

## Author contributions

### Credit authorship contribution statement

N. D. Zurita Cruz: methodology, formal analysis, investigation, writing.

A. Martín Ramírez: methodology, formal analysis, investigation, writing.

D. A. Rodríguez Serrano: resources, writing.

I. González Álvaro: resources, writing.

E. Roy Vallejo: resources, writing.

R. de la Cámara: resources, writing.

L. Fontán García-Rodrigo: methodology, formal analysis, writing.

L. Cardeñoso Domingo: conceptualization, investigation, methodology, writing, supervision.

All the authors have revised and approved the final version of the manuscript

## Acknowledgments

Special thanks to our patients and relatives for agreeing with the use of pseudonymized clinical data and surpluses of clinical samples to perform this study, to Manuel Gomez Gutierrez, PhD for his excellent editing assistance, and Ancor Sanz for assistance with statistical analysis.

PREINMUN-COVID group includes:

## Anesthesiology

Rosa Méndez, Julia Hernando, David Arribas.

## Hematology

Javier Ortiz, Isabel Iturrate, Rafael de la Cámara.

## Immunology

Francisco Sánchez-Madrid, Ildefonso Sánchez-Cerrillo, Pedro Martínez-Fleta, Celia López-Sanz, Ligia Gabrie, Luciana del Campo Guerola, Elena Fernández, Reyes Tejedor, Cecilia Muñoz, Aranzazu Alfranca. Ana Triguero-Martínez.

## Intensive Care Unit

Marta Chicot Llano, Pablo Patiño Haro, Marina Trigueros Genao, Begoña Quicios, Begoña González, Diego Aníbal Rodríguez Serrano.

## Internal Medicine-Infectious Diseases

Emilia Roy-Vallejo, Eduardo Sánchez, Fernando Moldenhauer, Marianela Ciudad, Ana Barrios, Beatriz Sánchez, Almudena Villa, Jesús Álvarez, Cristina Arévalo Román, José María Galván-Román, Jesús Sanz, Carmen Suárez Fernández.

## Microbiology

Nelly Daniela Zurita, Diego Domingo García, Teresa Alarcón Cavero, María Auxiliadora Semiglia Chong, Ainhoa Gutiérrez Cobos, María Diez Aguilar, Isabel García Arata, Laura Cardeñoso Domingo.

## Pneumology

Elena Ávalos, Celeste Marcos, Elena García Castillo, Ana Sánchez Azofra, Tamara Alonso, Carolina Cisneros, Claudia Valenzuela, Francisco Javier García Pérez, Rosa María Girón, Javier Aspa, Enrique Zamora, Mar Barrio Mayo, Rosalina Henares Espi, Olga Rajas, Joan B. Soriano, Julio Ancochea.

## References

1. Cheng ZJ, Shan J. 2020. 2019 Novel coronavirus: where we are and what we know. Infection 48:155–163.

2. WHO Coronavirus (COVID-19) Dashboard | WHO Coronavirus (COVID-19) Dashboard with Vaccination Data. https://covid19.who.int/. xRetrieved 30 June 2021.

3. Richardson S, Hirsch JS, Narasimhan M, Crawford JM, McGinn T, Davidson KW, Barnaby DP, Becker LB, Chelico JD, Cohen SL, Cookingham J, Coppa K, Diefenbach MA, Dominello AJ, Duer-Hefele J, Falzon L, Gitlin J, Hajizadeh N, Harvin TG, Hirschwerk DA, Kim EJ, Kozel ZM, Marrast LM, Mogavero JN, Osorio GA, Qiu M, Zanos TP. 2020. Presenting Characteristics, Comorbidities, and Outcomes among 5700 Patients Hospitalized with COVID-19 in the New York City Area. JAMA - Journal of the American Medical Association 323:2052–2059.

4. Asghar MS, Kazmi SJH, Khan NA, Akram M, Hassan M, Rasheed U, Khan SA. 2020. Poor Prognostic Biochemical Markers Predicting Fatalities Caused by COVID-19: A Retrospective Observational Study from a Developing Country. Cureus 12.

5. Zheng Z, Peng F, Xu B, Zhao J, Liu H, Peng J, Li Q, Jiang C, Zhou Y, Liu S, Ye C, Zhang P, Xing Y, Guo H, Tang W. 2020. Risk factors of critical & mortal COVID-19 cases: A systematic literature review and meta-analysis. Journal of Infection. W.B. Saunders Ltd https://doi.org/10.1016/j.jinf.2020.04.021.

6. Li F, Chen X, Zhao B, Qu Y, Chen Y, Xiong J, Feng Y, Men D, Huang Q, Liu Y, Yang B, Ding J, Li F. 2020. Clinical Infectious Diseases Detectable Serum Severe Acute Respiratory Syndrome Coronavirus 2 Viral Load (RNAemia) Is Closely Correlated with Drastically Elevated Interleukin 6 Level in Critically Ill Patients With Coronavirus Disease 2019. Clinical Infectious Diseases ® 71:1937–1979.

7. del Rio C, Malani PN. 2020. COVID-19 - New Insights on a Rapidly Changing Epidemic. JAMA - Journal of the American Medical Association. American Medical Association https://doi.org/10.1001/jama.2020.3072.

8. Lee S, Kim T, Lee E, Lee C, Kim H, Rhee H, Park SY, Son HJ, Yu S, Park JW, Choo EJ, Park S, Loeb M, Kim TH. 2020. Clinical Course and Molecular Viral Shedding among Asymptomatic and Symptomatic Patients with SARS-CoV-2 Infection in a Community Treatment Center in the Republic of Korea. JAMA Internal Medicine 180:1447–1452.

9. Tan C, Li S, Liang Y, Chen M, Liu J. 2020. SARS-CoV-2 viremia may predict rapid deterioration of COVID-19 patients. Brazilian Journal of Infectious Diseases 24:565–569.

10. Eberhardt KA, Meyer-Schwickerath C, Heger E, Knops E, Lehmann C, Rybniker J, Schommers P, Eichenauer DA, Kurth F, Ramharter M, Kaiser R, Holtick U, Klein F, Jung N, Cristanziano V di. 2020. RNAemia Corresponds to Disease Severity and Antibody Response in Hospitalized COVID-19 Patients. Viruses 12.

11. Hagman K, Hedenstierna M, Gille-Johnson P, Hammas B, Grabbe M, Dillner J, Ursing J. 2020. SARS-CoV-2 RNA in serum as predictor of severe outcome in COVID-19: a retrospective cohort study. Clinical Infectious Diseases https://doi.org/doi:10.1093/cid/ciaa1285.

12. Xu D, Zhou F, Sun W, Chen L, Lan L, Li H, Xiao F, Li Y, Kolachalama VB, Li Y, Wang X, Xu H. 2020. Relationship Between Serum Severe Acute Respiratory Syndrome Coronavirus 2 Nucleic Acid and Organ Damage in Coronavirus 2019 Patients: A Cohort Study. Clinical Infectious Diseases https://doi.org/10.1093/cid/ciaa1085.

13. Rodríguez-Serrano DA, Roy-Vallejo E, Zurita Cruz ND, Martín Ramírez A, Rodríguez-García SC, Arevalillo-Fernández N, Galván-Román JM, Fontán García-Rodrigo L, Vega-Piris L, Chicot Llano M, Arribas Méndez D, González de Marcos B, Hernando Santos J, Sánchez Azofra A, Ávalos Pérez-Urria E, Rodriguez-Cortes P, Esparcia L, Marcos-Jimenez A, Sánchez-Alonso S, Llorente I, Soriano J, Suárez Fernández C, García-Vicuña R, Ancochea J, Sanz J, Muñoz-Calleja C, de la Cámara R, Canabal Berlanga A, González-Álvaro I, Cardeñoso L. 2021. Detection of SARS-CoV-2 RNA in serum is associated with increased mortality risk in hospitalized COVID-19 patients. Scientific Reports 11:13134.

14. Bossuyt PM, Reitsma JB, Bruns DE, Gatsonis CA, Glasziou PP, Irwig L, Lijmer JG, Moher D, Rennie D, Korevaar DA, ré mie Cohen JF. 2015. STARD 2015: An Updated List of Essential Items for Reporting Diagnostic Accuracy Studies. British medical journal 351.

15. ThermoFisher Scientific. 2015. QuantStudio Design and Analysis desktop Software User Guide. (Pub. no. MAN0010408) Revision B.0.

16. Martín Ramírez A, Zurita Cruz ND, Gutiérrez-Cobos A, Rodríguez Serrano DA, González Álvaro I, Roy Vallejo E, Gómez de Frutos S, Fontán García-Rodrigo L, Cardeñoso Domingo L. 2022. Evaluation of two RT-PCR techniques for SARS-CoV-2 RNA detection in serum for microbiological diagnosis. Journal of Virological Methods 300:114411.

17. Heinrich F, Nentwich MF, Bibiza-Freiwald E, Nörz D, Roedl K, Christner M, Hoffmann A, Olearo F, Kluge S, Aepfelbacher M, Wichmann D, Lütgehetmann M, Pfefferle S. Blood SARS-CoV-2 RNA Load Predicts Outcome ° OFID ° 1 SARS-CoV-2 Blood RNA Load Predicts Outcome in Critically Ill COVID-19 Patients https://doi.org/10.1093/ofid/ofab509.

18. Chen L, Wang G, Long X, Hou H, Wei J, Cao Y, Tan J, Liu W, Huang L, Meng F, Huang L, Wang N, Zhao J, Huang G, Sun Z, Wang W, Zhou J. 2021. Dynamics of Blood Viral Load Is Strongly Associated with Clinical Outcomes in Coronavirus Disease 2019 (COVID-19) Patients. The Journal of Molecular Diagnostics 23:10–18.

19. Zheng S, Fan J, Yu F, Feng B, Lou B, Zou Q, Xie G, Lin S, Wang R, Yang X, Chen W, Wang Q, Zhang D, Liu Y, Gong R, Ma Z, Lu S, Xiao Y, Gu Y, Zhang J, Yao H, Xu K, Lu X, Wei G, Zhou J, Fang Q, Cai H, Qiu Y, Sheng J, Chen Y, Liang T. 2020. Viral load dynamics and disease severity in patients infected with SARS-CoV-2 in Zhejiang province, China, January-March 2020: retrospective cohort study. BMJ 369.

20. Agarwal J, Singh V, Garg J, Karoli R, Tiwari S, Naqvi S, Das A, Sen M. 2021. RNAemia and Clinical Outcome in COVID-19 Patients. Journal of Microbiology and Infectious Diseases J Microbiol Infect Dis 11:116–123.

21. Colagrossi L, Antonello M, Renica S, Merli M, Matarazzo E, Travi G, Vecchi M, Colombo J, Muscatello A, Grasselli G, Molteni SN, Scaravilli V, Cattaneo E, Fanti D, Vismara C, Bandera A, Gori A, Puoti M, Cento V, Alteri C, Perno CF. SARS-CoV-2 RNA in plasma samples of COVID-19 affected individuals: a cross-sectional proof-of-concept study https://doi.org/10.1186/s12879-021-05886-2.

22. Hogan CA, Stevens BA, Sahoo MK, Huang C, Garamani N, Gombar S, Yamamoto F, Murugesan K, Kurzer J, Zehnder J, Pinsky BA. Clinical Infectious Diseases High Frequency of SARS-CoV-2 RNAemia and Association with Severe Disease https://doi.org/10.1093/cid/ciaa1054.

23. Fajnzylber J, Regan J, Coxen K, Corry H, Wong C, Rosenthal A, Worrall D, Giguel F, Piechocka-Trocha A, Atyeo C, Fischinger S, Chan A, Flaherty KT, Hall K, Dougan M, Ryan ET, Gillespie E, Chishti R, Li Y, Jilg N, Hanidziar D, Baron RM, Baden L, Tsibris AM, Armstrong KA, Kuritzkes DR, Alter G, Walker BD, Yu X, Li JZ. 2020. SARS-CoV-2 viral load is associated with increased disease severity and mortality. Nature communications 11:5493-.

24. Jacobs JL, Mellors JW. Detection of Severe Acute Respiratory Syndrome Coronavirus 2 (SARS-CoV-2) RNA in Blood of Patients with Coronavirus Disease 2019 (COVID-19): What Does It Mean? https://doi.org/10.1093/cid/ciaa1316.

25. Santos Bravo M, Nicolás D, Berengua C, Fernandez M, Hurtado JC, Tortajada M, Barroso S, Vilella A, Mosquera MM, Vila J, Marcos MA. 2021. The Journal of Infectious Diseases Severe Acute Respiratory Syndrome Coronavirus 2 Normalized Viral Loads and Subgenomic RNA Detection as Tools for Improving Clinical Decision Making and Work Reincorporation. The Journal of Infectious Diseases ® 224:1325–1357.

26. Berger A, Braner J, Doerr HW, Weber B. 1998. Quantification of Viral Load: Clinical Relevance for Human Immunodeficiency Virus, Hepatitis B Virus and Hepatitis C Virus Infection. Intervirology 41:24–34.

27. Pawlotsky JM, Bouvier-Alias M, Hezode C, Darthuy F, Remire J, Dhumeaux D. 2000. Standardization of Hepatitis C Virus RNA Quantification. Hepatology 32:654–659.

28. Peter JB, Sevall JS. 2004. Molecular-Based Methods for Quantifying HIV Viral Load. https://home.liebertpub.com/apc 18:75–79.

29. Guiver M, Fox AJ, Mutton K, Mogulkoc N, Egan J. 2001. Evaluation of CMV viral load using TaqMan CMV quantitative PCR and comparison with CMV antigenemia in heart and lung transplant recipients. Transplantation 71:1609–1615.

30. Xu D, Zhou F, Sun W, Chen L, Lan L, Li H, Xiao F, Li Y, Kolachalama VB, Li Y, Wang X, Xu H. 2021. Relationship Between Serum Severe Acute Respiratory Syndrome Coronavirus 2 Nucleic Acid and Organ Damage in Coronavirus 2019 Patients: A Cohort Study. Clinical Infectious Diseases 73:68–75.

31. Veyer D, Kernéis S, Poulet G, Wack M, Robillard N, Taly V, L’Honneur A-S, Rozenberg F, Laurent-Puig P, Bélec L, Hadjadj J, Terrier B, Péré H. 2020. Highly Sensitive Quantification of Plasma Severe Acute Respiratory Syndrome Coronavirus 2 RNA Sheds Light on its Potential Clinical Value. Clinical Infectious Diseases https://doi.org/10.1093/cid/ciaa1196.

32. Jacobs JL, Bain W, Naqvi A, Staines B, Castanha PMS, Yang H, Boltz VF, Barratt-Boyes S, Marques ETA, Mitchell SL, Methé B, Olonisakin TF, Haidar G, Burke TW, Petzold E, Denny T, Woods CW, Mcverry BJ, Lee JS, Watkins SC, St Croix CM, Morris A, Kearney MF, Ladinsky MS, Bjorkman PJ, Kitsios G, Mellors JW. SARS-CoV-2 Viremia is Associated with COVID-19 Severity and Predicts Clinical Outcomes https://doi.org/10.1093/cid/ciab686/6347519.

33. Peña KB, Riu F, Gumà J, Guilarte C, Pique B, Hernandez A, Àvila A, Parra S, Castro A, Rovira C, Cueto P, Vallverdu I, Parada D. 2021. Study of the Plasma and Buffy Coat in Patients with SARS-CoV-2 Infection-A Preliminary Report Academic Editors: Natalia Rakislova and Jaume Ordi https://doi.org/10.3390/pathogens10070805.

